# Transmission of hepatitis E virus in Bangladesh: A phylogenetic analysis of viral sequences from a nation-wide hospital-based acute jaundice surveillance program, 2014-2017

**DOI:** 10.1101/2025.01.09.25320246

**Authors:** Md Mobarok Hossain, Amy Dighe, Repon C Paul, Arifa Nazneen, Lilia Ganova-Raeva, Saleem Kamili, Firdausi Qadri, Andrew S. Azman, Emily S. Gurley, Mustafizur Rahman

## Abstract

**Background:** Hepatitis E virus (HEV) genotype 1 is a major cause of acute jaundice in Bangladesh, yet the transmission dynamics and genetic diversity of this virus remains inadequately characterized. This study aims to elucidate the genetic landscape and transmission patterns of HEV infection in Bangladesh through phylogenetic analysis of viral sequences obtained from a nation-wide surveillance program.

**Methodology/Principal Findings:** We analyzed 104 partial HEV open reading frame 1 (ORF-1) sequences collected from acute jaundice patients admitted to six tertiary hospitals across Bangladesh during December 2014– September 2017. Phylogenetic trees were constructed using maximum likelihood methods, and Bayesian clustering was employed to assess genetic diversity and transmission patterns. All sequences were identified as HEV genotype 1 (HEV-1), with 10 sequences predominantly collected in 2017 classified as subtype 1g, forming a distinct cluster. A lack of geographic clustering across the sequences suggests widespread transmission across the country rather than geographically distinct transmission networks. Of the 104 sequenced cases, 5 (5%) were associated with fatal outcomes, although these sequences did not cluster phylogenetically.

**Conclusions/Significance:** This phylogenetic analysis provides evidence of widespread transmission of HEV-1 across Bangladesh, with a reduction in genetic diversity in 2017 suggesting the potential emergence of a dominant viral cluster around that time. Given the paucity of clinical surveillance of HEV, genomics may provide new insights into unobserved aspects of the transmission of the virus locally and globally.

**Author summary:** Hepatitis E virus genotype 1 (HEV-1) is a major cause of acute jaundice in Bangladesh, but many cases go unreported, and the virus’s transmission patterns are not well understood. In the absence of reliable clinical surveillance data, analyzing the genetic diversity amongst viral samples isolated from infected people can help us infer transmission patterns. To better understand how HEV-1 circulates in Bangladesh we studied 104 HEV-1 genetic sequences isolated from acute jaundice patients at 6 hospitals across the country between 2014-17 and inferred how the sequences most likely relate to each other based on their genetic similarities. We found that the viral sequences did not cluster by region in which the patient lived but rather genetically similar viruses were isolated in geographically distant regions suggesting widespread transmission across the country. Our results captured a reduction in genetic diversity in 2017 with many sequences isolated that year forming a distinct genetically similar group suggesting a potential shift in the viral population or a significant outbreak event around that time. Given the scarcity of clinical surveillance of HEV-1, such insights into unobserved aspects of transmission have the potential to improve our understanding of HEV-1 epidemiology.

## Introduction

Over the last decade phylogenetics have refined our understanding of the transmission of numerous viral pathogens, yet there has been relatively little application of these approaches to hepatitis E virus (HEV) genotype 1 (HEV-1). HEV-1 is spread from person-to-person via contaminated water and cause large recurring outbreaks of acute jaundice, as well as sporadic cases, in endemic regions across South Asia and parts of Africa (Al-Shimari et al., 2023; Koyuncu et al., 2021; Rein et al., 2012; Stanaway et al., 2016). Infection usually causes self-limiting disease, but carries a risk of acute liver failure which is especially high for pregnant women among whom the measured case fatality rate (CFR) is from 0-65% (Amanya et al., 2017; Kamar et al., 2012; Koyuncu et al., 2021). Despite presenting an important public health problem, the scarcity of diagnostics and underreporting (Koyuncu et al., 2021) implies that the transmission dynamics of HEV-1 are not well understood both at the micro- and macroscales (i.e., regional/global). Additionally, little is known about the genetic differences between outbreak and circulating strains or how viral genetics affect the severity of disease.

In Bangladesh, HEV-1 is a major cause of clinically attended acute jaundice and fulminant hepatitis (Mamun-Al-Mahtab et al., 2009; Paul et al., 2020; Sheikh et al., 2002). The first outbreak of non-A non-B hepatitis in Bangladesh was reported in Dhaka in 1987 (Hlady et al., 1990), but HEV is likely to have been present in the country much earlier than this date. On top of the sporadic cases and small clusters which occur across the country throughout the year (Hoa et al., 2021; Paul et al., 2020; Sazzad, Labrique, et al., 2017; Sazzad, Luby, et al., 2017), large outbreaks frequently cause significant morbidity, mortality, and disruption to people and their communities. Over the past 20 years, seven major HEV outbreaks have been reported in the peer reviewed literature, all of which have occurred in urban areas, with the first two in Dhaka in 2004 and 2008-9, two in Rajshahi City Corporation in 2010 and 2017, one in Noakhali in 2013, and two in Chattogram City in 2012 and 2018 (Aziz et al., 2022; Baki et al., 2021; Biswas et al., 2020; Gurley et al., 2014; Haque et al., 2015; Harun-Or-Rashid et al., 2013; Owada et al., 2022; Rahman et al., 2018). The majority of these outbreaks have been traced back to fecal contamination of municipal water supplies, but outside of this mode, our collective understanding of the transmission of HEV-1 across Bangladesh and over time remains nascent.

Viral sequencing of HEV-1 in Bangladesh has been limited to samples from two of the major outbreaks and from sporadic cases seen by a single hospital site and a private laboratory in Dhaka (Baki et al., 2021; Biswas et al., 2020; Harun-Or-Rashid et al., 2013; Hoa et al., 2021). The two viral isolates sampled during the 2010 outbreak in Rajshahi were sequenced and classified as genotype 1a (Harun-Or-Rashid et al., 2013). When further phylogenetic analysis was conducted, these two sequences clustered with the 21 other Bangladeshi sequences from sporadic cases in Dhaka between 2013 and 2015 (Hoa et al., 2021). Although this could suggest that viruses that cause sporadic cases are also capable of causing outbreaks, it is difficult to make reliable inferences about transmission dynamics without comparison with a larger number of sequences from Bangladesh and neighboring countries. All sequences sampled from the 2018 outbreak in Chittagong were deemed genotype 1f (Baki et al., 2021; Biswas et al., 2020), and the similarity between sequences from the outbreak was very high at 99.8% (Baki et al., 2021). The relationship with previous outbreak strains and with circulating strains is not known. More sequences from a wider geographic area are necessary to better characterize the spread of HEV infection across Bangladesh and over time, and to understand the contribution of viral genetics to outbreak emergence and disease severity.

We use phylogenetic analysis of 104 partial viral genome sequences, collected as part of a nation-wide hospital-based acute jaundice surveillance program during 2014 to 2017 (Paul et al., 2020), to gain insight into the transmission of HEV over time and across geographies in Bangladesh.

## Methods

### Sample and data collection

The viral sequences analyzed were collected from acute jaundice patients identified through a hospital-based surveillance study led by icddr,b, Dhaka, with the primary aim of estimating the burden of HEV infection in hospitalized patients with acute jaundice in Bangladesh. Surveillance was conducted from December 2014 to September 2017 at six tertiary hospitals spread across five of the seven administrative divisions of Bangladesh as defined at the time of the study, using enrolment criteria described previously (Paul et al., 2020). In brief, the study team in each hospital attempted to enroll all patients with acute jaundice admitted to the adult medicine and obstetrics and gynecology wards (serving patients aged ≥14 years). Acute jaundice was defined as a new onset of yellow eyes or yellow skin that began less than three months prior to hospital admission. At enrollment, a 5 ml blood sample was collected from each acute jaundice patient, along with basic demographic and clinical information. The sera were tested for IgM and IgG antibodies to HEV (anti-HEV) using enzyme-linked immunosorbent assay (ELISA) kits (Beijing Wantai Biologic Pharmacy Enterprise Co., Ltd, Beijing, China). Acute jaundice patients were followed during their hospital stay, as well as three months after discharge (or three months after the expected delivery date in the case of pregnancy) to capture outcomes of infection.

In total, 1925 acute jaundice patients were identified through the surveillance program, 661 of which had anti-HEV IgM indicative of acute infection. All samples taken from patients with symptom onset within three weeks of blood draw, plus a random sample of 20% of the remaining IgM positive patients, 20% of anti-HEV IgG positive patients, and 20% of patients who were both IgM and IgG negative, were sent to the Division of Viral Hepatitis Laboratory of the US Centers for Disease Control and Prevention (CDC), Atlanta, GA, USA for RNA testing. RNA testing was performed using a quantitative real-time reverse transcriptase polymerase chain reaction (PCR) assay targeting a 69-bp fragment of open reading frame 3 (ORF-3) of the HEV genome. This assay has been shown to be capable of detecting HEV genotypes 1–3 with a limit of detection of 25 IU/ml (Jothikumar et al., 2006). Of the 138 IgM positive samples tested, 131 had evidence of HEV RNA.

### Sequencing

ORF-1 was chosen as a sequencing target for samples with HEV RNA. This region was chosen in keeping with historical use by the CDC for genotyping HEV, as it includes conserved regions allowing for the identification of genotype but also includes hypervariable regions that allow for distinguishing between viruses over relatively short spatial and temporal scales (Smith, et all, 2013). There was not sufficient material to attempt whole genome sequencing for any of the 131 samples, as the sensitivity of the PCR protocol is not equal across all genomic regions. Consequently, to allow for phylogenetic analysis, ORF1 was successfully amplified from 104 HEV RNA positive isolates.

### Phylogenetic analysis

We conducted a phylogenetic analysis using 104 study sequences of HEV partial ORF1, along with 38 HEV-1 sequences obtained from GenBank. These included 23 sequences from Bangladesh (accession numbers: MH991993-MH992013, AB369397 and AB720035), and 15 sequences isolated outside of Bangladesh. Additionally, we used 5 HEV-2,3 and 4 sequences (accession numbers: KX578717, AF082093, AF279122, AF264009 and AF264010) to root the phylogenetic tree. We aligned the sequences using the multiple sequence alignment tool, MAFFT (Katoh et al., 2009), and derived a single nucleotide polymorphism (SNP) matrix from the alignment file using the software snp-dists (Seemann, 2018).

We built maximum likelihood trees (MLT) from the alignments using RAxML v8.2.12 (Stamatakis, 2014), applying the Generalized Time-Reversible model with a Gamma distribution to account for site-specific rate variation (GTRGAMMA in RAxML). We calculated support for the MLT using 1000 bootstrap pseudo-analyses of the alignment. We then implemented a hierarchical Bayesian clustering algorithm in Fastbaps v1.0 (Fast Hierarchical Bayesian Analysis of Population Structure) (Tonkin-Hill et al., 2019) in R v3.5.3 using the ape v5.3, ggplot2 v3.1.1, and ggtree v2.4.1 packages to cluster the alignment sequences (Paradis & Schliep, 2019; R Core Team, 2021; Wickham, 2016; Yu et al., 2017). We visualized and annotated the phylogenetic trees using iTOL v6 (Letunic & Bork, 2021). Finally, we attempted to classify the study sequences into subtypes using the RIVM subtyping tool (RIVM, 2020; Smith et al., 2020).

### Ethics statement

Consent to participate in the study was sought from the patients or, in the case of critically ill patients, from their guardians. For patients over 17 years old, written informed consent was obtained. For those aged between 14 and 17 years, both written assent from the patients and written consent from their parents or guardians were taken. The study protocol was reviewed and approved by the icddr,b institutional review board.

## Results

We sequenced the methyltransferase gene (216 bp in ORF-1) of HEV from 104 acute jaundice patients admitted to six tertiary hospitals across Bangladesh from December 2014 to September 2017 (GenBank accession numbers PQ431073-PQ431176) (Figure 1). Sequences were isolated from patients across all six hospitals throughout the study period, representing individuals who lived in every division of Bangladesh (Figure 1B & C). The proportion of IgM positive samples that were sent for PCR testing was relatively consistent across sites ranging from 24-42%, with the proportion of those successfully sequenced ranging from 42-67%. The increased number of sequences collected in 2016 (Figure 1C) is proportional to an increase in IgM positive acute jaundice cases enrolled compared to 2015. Sequencing efforts were consistent across the years, with 1 of the 2 IgM positive cases sequenced in December 2014, 19% of IgM positive cases sequenced in both 2015 (n = 28/145) and 2016 (n = 50/259), and 10% (n = 25/255) in 2017.

**Figure 1.**
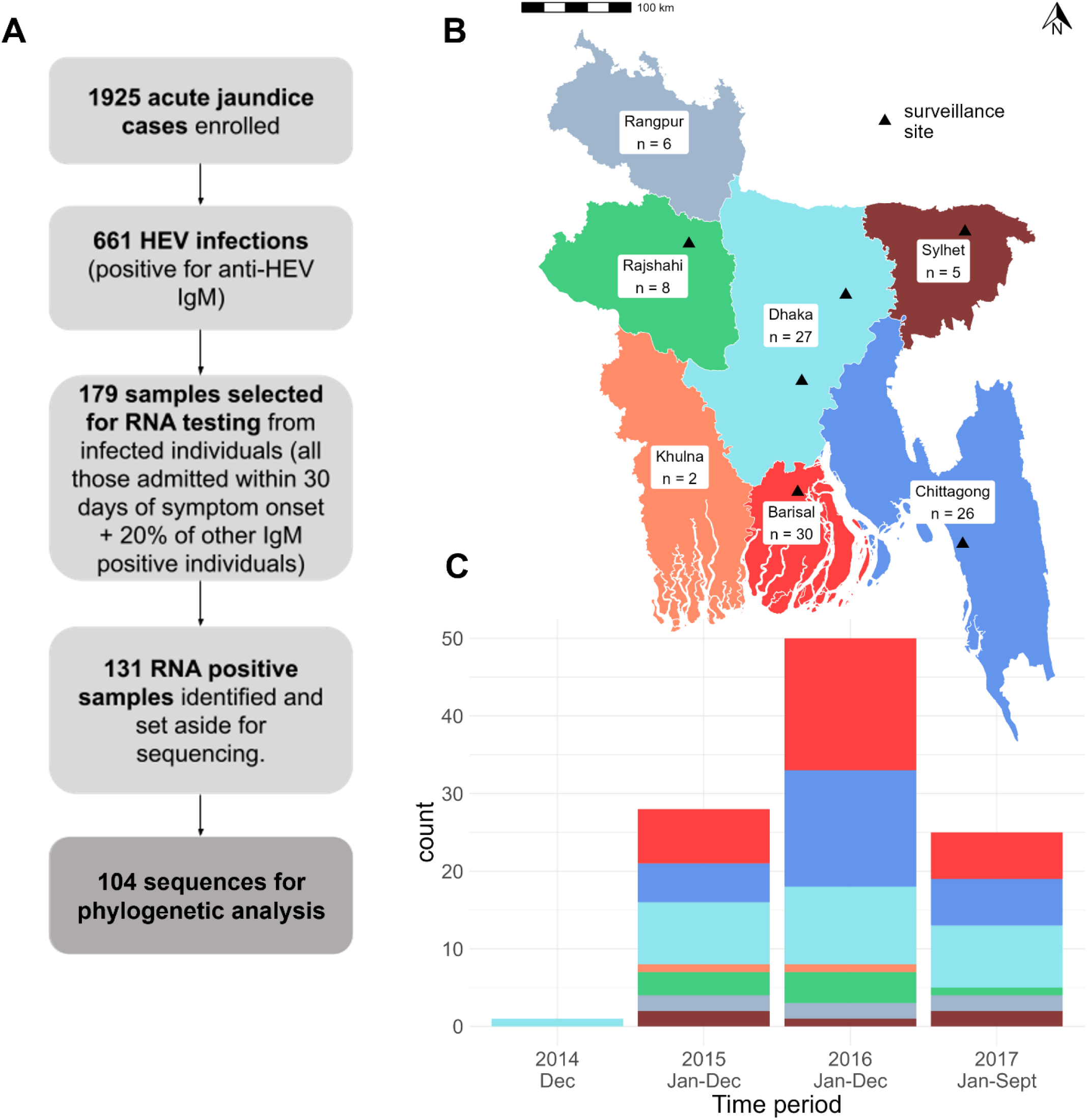
**A**. The selection of samples for sequencing. **B**. The distribution of sequences across the divisions of Bangladesh where HEV infected patients lived. Please note that administrative boundaries reflect the official boundaries in 2014 when the survey was designed, rather than the current boundaries. **C**. The distribution of sequences collected by year and by division where the patient lived. The location of the 6 tertiary hospitals are shown as black triangles.

All the HEV sequences belonged to genotype 1. The mean genetic distance between isolates was approximately 6.6 single nucleotide polymorphisms (SNPs), corresponding to an average sequence identity of 96.9%. Only 10 of the sequences could be reliably sub-typed, all of which were classified as subtype 1g which was defined in 2017 after a HEV-1 strain isolated in Mongolia in 2015 was found to be genetically distinct from pre-defined subtypes 1a-1f and has subsequently been isolated from patients in India, Pakistan, Japan, UK, and France (Tsatsralt-Od et al., 2018; Nishizawa et al., 2017; Smith et al., 2020). The 1g sequences from our study formed their own cluster C4 shown in brown on the inner ring (1) in the phylogenetic tree (Figure 2A) together with the 1g sequence collected in France which was available on GenBank (accession number: MN401238). We observed some temporal clustering, with all but one of the C4 sequences isolated from patients admitted in 2017, but these were not geographically clustered; rather they came from patients living across four different divisions (Barisal, Sylhet, Dhaka and Rajshahi) (Figure 2A, Figure S1). Indeed, more generally, based on the information in the methyltransferase gene, the sequences did not cluster geographically but were well mixed across the country (Figure 2A). The phylogenetic analysis revealed that the majority of the major branches in the tree exhibited strong bootstrap support, with values ranging from 75% to 100% (C1-C4 cluster). This high level of bootstrap support indicates that the grouping of sequences within these branches is highly reliable and reflects robust phylogenetic relationships. However, some of the smaller branches within cluster C5 displayed lower bootstrap values, suggesting that the phylogenetic relationships in these areas are less certain and may be subject to variability across different bootstrap replicates (Figure 2A).

**Figure 2.**
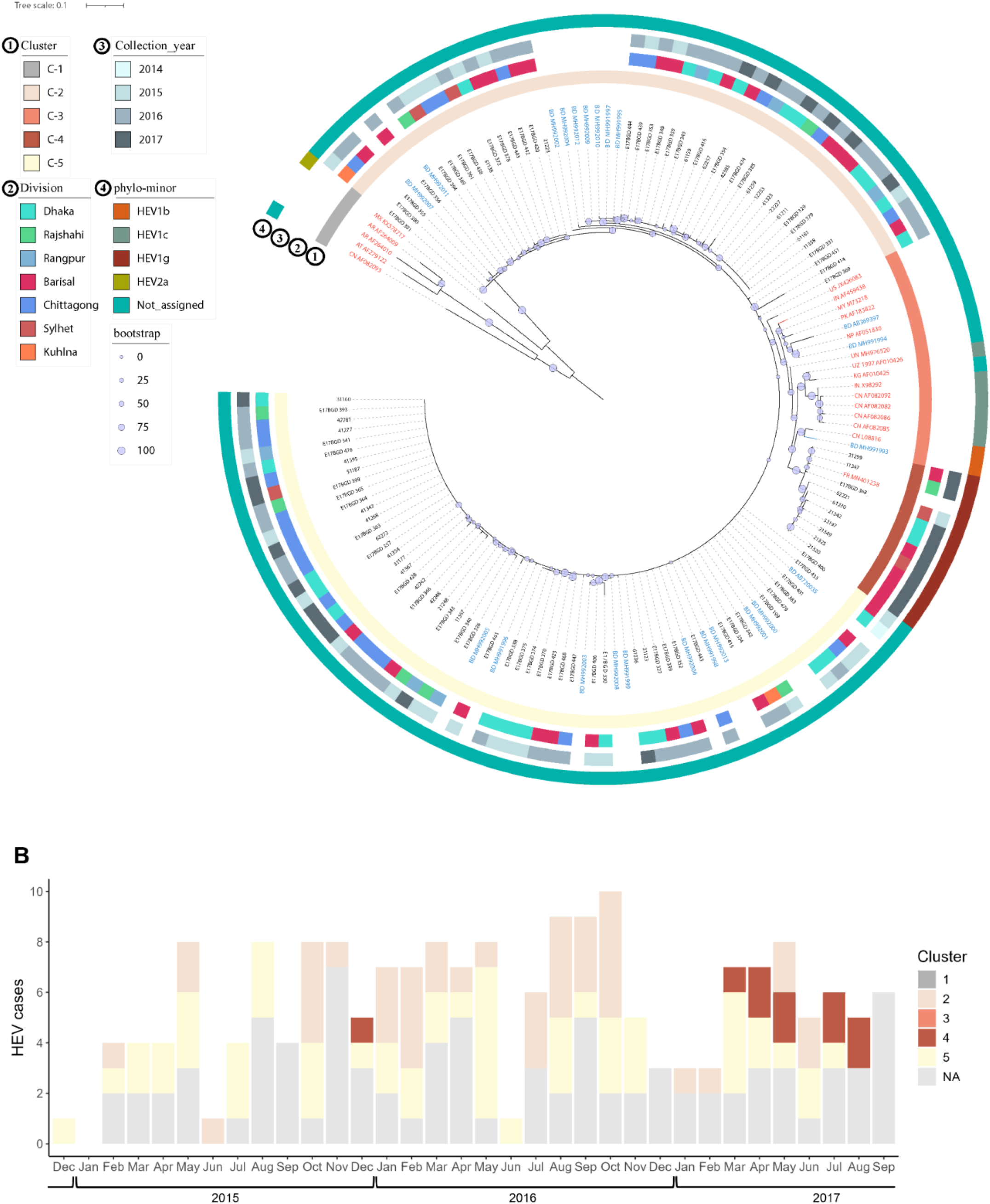
**A**. Phylogenetic relationship among hepatitis E virus (HEV) strains from Bangladesh and reference strains available in GenBank, based on a 216-nt sequence in open reading frame 1 (ORF-1). Each branch is labeled with the GenBank accession number, BAPs cluster, division of sample collection area, collection date, and the identified phylo-minor genotype. The phylo-minor genotype was determined using the hepatitis E virus RIVM subtyping tool (RIVM, 2020; Smith et al., 2020). Scale bars represent nucleotide substitutions per site. Sequences from this study are labeled in black text while GenBank sequences are labeled in red text if collected outside of Bangladesh and blue if collected in Bangladesh. Bootstrap values for branches are indicated using gray circles and represent how many times out of 100 that the same branch is observed when the phylogenetic tree is regenerated on a resampled set of the data. **B**. HEV IgM seropositive acute jaundice cases admitted at 6 tertiary hospitals across Bangladesh colored by phylogenetic cluster as defined by fastBAPS and based on the methyltransferase gene. Cases without a sequenced sample are indicated in light gray. Clusters 1 and 3 are absent from the curve as 1 refers to non HEV sequences used to root the tree, and members of 3 were all from GenBank and not collected during our study.

The methyl transferase sequences collected by previous studies in Bangladesh were dispersed throughout our study sequences in the phylogenetic tree (indicated by the blue text in Figure 2A). These included 21 sequences collected from sporadic hepatitis E cases in Dhaka during 2013–2015, 1 sequence from the 2010 outbreak in Rajshahi (BD_AB720035), and 1 sequence isolated in 2002 from a patient in Japan who had traveled to Bangladesh (BD_AB369397). The HEV-1 sequences from outside of Bangladesh (red text in Figure 2A) tended to group together in cluster 3, along with some of the sequences from the Dhaka study. The sequences from outside of Bangladesh also tended to be from earlier years, with known dates of collection spanning 1986-2013.

Of the 104 study sequences, 5 (5%) were associated with fatal cases of acute jaundice. These 5 sequences did not cluster together phylogenetically, and were spread across the two largest clusters (C2 and C5), as defined using a Bayesian clustering algorithm, BAPS (Tonkin-Hill et al., 2019).

## Discussion

For diseases like hepatitis E, for which clinical surveillance data is very limited in endemic regions, genomic analyses can provide valuable insights into transmission patterns and inform public health interventions. Over the past decade viral sequencing and phylogenetics have transformed our understanding of the transmission of many pathogens that pose a risk to public health. However, whilst non-zoonotic HEV-1 has been increasingly recognized as an important public health problem in parts of Africa and Asia, there have been relatively few published sequences or phylogenetic analyses of the virus. We were able to sequence 104 partial HEV-1 genomes from hospitalized patients enrolled in an acute jaundice surveillance study conducted across Bangladesh. Although constrained by the information stored in a relatively short region of the viral genome, phylogenetic analysis of these sequences along with existing publicly available sequences, provided several important insights into the transmission of HEV-1 geographically and over time within the country.

All the partial genomes isolated and sequenced in the study belonged to genotype 1 suggesting that despite the presence of HEV in animals in Bangladesh (Haider et al., 2017), zoonotic transmission of HEV-3 or 4 is not making a significant contribution to clinical hepatitis E cases. To some extent sequences clustered by time of isolation, with a number of methyltransferase sequences isolated in 2017 forming a distinct genetically similar group of subtype-1g sequences forming their own cluster within the other study sequences. We did not, however, see any notable clustering of sequences by the administrative division where the patient lived, which would suggest that transmission of viruses occurred widely across the whole country rather than being highly structured into localized transmission networks. It is important to acknowledge that this finding only takes into account a relatively short region of the genome and whole genome sequencing would be required for more robust analysis of clusters over space and time, and a greater understanding of transmission dynamics across the country and internationally between endemic regions.

During the study period there was only 1 major outbreak of HEV infection reported in the literature, which occurred in Rajshahi in 2017 (Aziz et al., 2022) but little detail was published and there were no known sequencing efforts. Phylogenetic analysis of isolates from outbreaks have found them to be highly genetically similar or identical to one another (Baki et al., 2021). We did not see any clusters of sequences from Rajshahi and did not capture an increase in cases reported in Bogra during 2017 so we assume that most if not all study sequences represent the sequence diversity of viruses contributing to ongoing sporadic transmission or smaller unreported local outbreaks. Methyltransferase sequences were only publicly available for a single outbreak (the 2010 outbreak in Rajshahi) so it was not possible to provide a comprehensive genetic comparison of sequences from so-called sporadic cases and outbreak strains within Bangladesh. Future efforts to conduct phylogenetic analysis of outbreak and endemic viruses could identify whether or not there are viral genetic factors that contribute to the occurrence of outbreaks. Continued genetic surveillance in parallel to epidemiological surveillance over time could allow for the exploration of how viral genetics affect outbreak risk in the context of host immunity and past infection history, which could be particularly important if considering vaccine introduction.

There are several limitations of our analysis. First, we analyzed a relatively short sequence (216 bp) because we only had access to a small proportion of the genetic information held in the HEV genome with which to infer genetic distance between the viruses, meaning there is a chance that the structure of the phylogenetic tree could have looked different if it was based on information from the whole genome. Likely due to the length of the sequence, there was not a clear correlation between time of isolation and genetic distance so we could not justify use of a molecular clock model to resolve the phylogeny over time, though we were able to gain some insight into temporal dynamics by looking at how year of admission was distributed across the phylogenetic tree. In addition, inconsistency in the sub-genomic region chosen for sequencing in different studies meant we could only include a small proportion of publicly available HEV sequences in our analysis. Finally, our sampled population were hospitalized acute jaundice patients, meaning all sequences came from relatively severe cases of hepatitis E and therefore may not fully capture the genetic diversity of HEV infection in the general population. The absence of sequences from mild cases and subclinical infections limited our ability to look at the association between viral genetics and severity, however, we did not see genetic clustering of the five sequences associated with fatal cases of acute jaundice.

In summary, whilst constrained by short sequence length, the phylogenetic analysis of 104 HEV-1 partial genome sequences from acute jaundice patients with sporadic HEV infections seems to capture a reduction in genetic diversity in 2017, with increased dominance of a genetically similar cluster of sub-type 1g viruses in several divisions across Bangladesh. This suggests a potential shift in the viral population or a significant outbreak event that influenced the virus’s genetic landscape. The lack of evidence of geographic clustering of the sequences suggests widespread transmission of HEV across Bangladesh rather than localized circulation. Future studies conducting whole genome sequencing using Next Generation shotgun library approach or focusing on a consistent, larger sub-genomic target are necessary for more robust analysis (Purdy, et all., 2017) of the transmission dynamics of HEV infection over time and geographically within Bangladesh and other endemic areas.

## Data Availability

The sequences data generated and used in this work have been made available on GenBank (https://www.ncbi.nlm.nih.gov/genbank/) under accession numbers PQ431073-PQ431176.

## Acknowledgements

We thank all the participants in the surveillance study for taking part in this research. We thank the field and laboratory staff for their dedicated work. We thank Dr. M. Purdy for his expertise and advice.

## Disclaimer

**The findings and conclusions in this report do not necessarily reflect the official position of the Centers for Disease Control and Prevention, or the authors’ affiliated institutions. Use of trade names and commercial sources is for identification only and does not imply endorsement by the Centers for Disease Control and Prevention, the Public Health Service, or the US Department of Health and Human Services**.

## Data reporting

The sequences generated have been made available on GenBank under accession numbers PQ431073-PQ431176.

